# CMR Diffusion Tensor Imaging Provides Novel Imaging Markers of Adverse Myocardial Remodeling in Aortic Stenosis

**DOI:** 10.1101/2020.08.17.20176297

**Authors:** Alexander Gotschy, Constantin von Deuster, Lucas Weber, Mareike Gastl, Martin O. Schmiady, Robbert J. H. van Gorkum, Johanna Stimm, Jochen von Spiczak, Robert Manka, Sebastian Kozerke, Christian T. Stoeck

## Abstract

**Objectives:** This study sought to determine microstructural cardiac remodeling in aortic stenosis (AS) and its reversibility following valve replacement using cardiovascular magnetic resonance (CMR) diffusion tensor imaging (DTI).

**Background:** Myocardial involvement in AS, such as focal and diffuse fibrosis is associated with worse outcome, even after timely aortic valve replacement (AVR). Alterations of myofiber architecture and myocardial diffusion may precede fibrosis, but its extent and reversibility after AVR are unknown.

**Methods:** Patients with isolated severe AS (n = 21, 62% male; mean age 75 years) and sex-matched senior control subjects underwent prospective CMR DTI. Changes in the DTI parameters: mean diffusivity (MD), fractional anisotropy (FA) as well as helix angle (HA) and absolute E2A sheet angle (E2A) were quantified and compared with native T1 and extracellular volume (ECV) as standard CMR markers of myocardial fibrosis. Six months after AVR eleven patients were scheduled for a follow-up CMR.

**Results:** In AS patients, significantly elevated MD (p = 0.002) and reduced FA (p< 0.001) were measured when compared to controls. Myocyte aggregate orientation exhibited a steeper transmural HA slope (p< 0.001) and increased absolute E2A sheet angle (p< 0.001) in AS. Six months post AVR, the HA slope (p< 0.001) was reduced to the level of healthy controls and MD (p = 0.014), FA (p = 0.011) and E2A (p = 0.003) showed a significant regression towards normal values. In contrast, native T1 was similar in AS and controls and did not change significantly after AVR. ECV showed a non-significant trend (p = 0.16) to higher values after AVR.

**Conclusion:** In patients with severe aortic stenosis, CMR DTI provides a set of parameters that identifies structural and diffusion abnormalities, which are largely reversible after AVR. DTI parameters showed proportionally greater changes in response to AS and AVR compared to metrics of myocardial fibrosis and may, therefore, aid risk stratification in earlier stages of severe AS.

**Condensed Abstract:** CMR diffusion tensor imaging (DTI) is a novel, noninvasive technique that allows for the assessment of myocardial microstructure in diseased hearts. We used CMR DTI to investigate myocardial involvement in patients with aortic stenosis (AS) before and after aortic valve replacement (AVR). Measures of tissue diffusion as well as parameters of myocyte orientation were significantly altered by AS and showed a clear trend towards normalization after AVR. Conventional markers of myocardial fibrosis (native T1 & extracellular volume) did not change significantly after AVR. Therefore, CMR DTI may unlock a new level of detail for phenotyping myocardial involvement in AS with potential value for improved risk stratification by visualizing earlier stages of adverse remodeling.

## Introduction

Aortic stenosis (AS) affects up to 4.6% of the population older than 75 years of age (1) and is the most common cause for valve intervention in the Western world (2). Despite recent advances in surgical and transcatheter aortic valve replacement (AVR), AS patients have shorter life expectancy compared to the general population, even after timely intervention (3). While current guidelines focus on the hemodynamic assessment at the level of the valve for the timing of intervention (4), the prognostic role of adverse myocardial response becomes increasingly evident (5). Myocardial fibrosis has been identified as a key driver in the transition from afterload induced physiological adaption to irreversible remodeling and decompensation (6). Focal myocardial fibrosis, identified by cardiac magnetic resonance (CMR) based late gadolinium enhancement (LGE) has been shown to be an independent predictor of mortality in AS patients (7,8) with a hazard ratio of 2.5 for all-cause mortality. Likewise, measures of diffuse fibrosis such as native T1 and extracellular volume (ECV) were found to be associated with outcome in AS (9,10). After AVR, the progression of focal fibrosis can be stopped and diffuse fibrosis is partially reversible (11,12). However, if present before AVR, both diffuse and focal fibrosis predict significantly increased late mortality after AVR. Compared to patients without evidence of myocardial fibrosis, all-cause mortality of patients with LGE is 2-fold higher and even up to 3-fold higher in patients with significantly elevated ECV over 3–4 years of follow-up after AVR (13,14). Hence, earlier markers of adverse myocardial remodeling may improve patient selection before irreversible damage occurs, in particular in light of increasing evidence for intervention in asymptomatic severe AS (15).

Beyond imaging of the passive, fibrotic compartment of the left ventricle (LV), CMR diffusion tensor imaging (DTI) offers a novel approach for phenotyping myocyte response to adverse stimuli. It allows for the assessment of cardiomyocyte aggregate orientation in three dimensions including the quantification of myocardial diffusion metrics (16). In patients with cardiac amyloidosis, hypertrophic and dilated cardiomyopathy, characteristic alterations of those parameters have been described and partially correlated with functional LV impairment (17–20).

In the present study, we investigated CMR DTI in patients with severe AS. In particular, we assessed alterations of myocardial microstructure due to AS compared with healthy controls and investigated the reversibility of changes in the DTI parameters after AVR in comparison to markers of myocardial fibrosis.

## Methods

### Study Design

For this prospective, observational study, 21 patients with severe AS and 10 healthy controls were recruited between June 2016 and January 2020 in a single tertiary referral heart center at the University Hospital Zurich, Switzerland. Inclusion criteria were adult patients with severe AS as determined by echocardiography (aortic valve area < 1 cm^2^, indexed aortic valve area < 0.6 cm^2^/m^2^ or mean pressure gradient > 40 mm Hg) in the presence of normal LV ejection fraction (LVEF > 50%). Exclusion criteria were previous myocardial infarction, obstructive coronary artery disease on angiography, any cardiomyopathy, any moderate or severe valve disease besides AS, kidney failure with estimated glomerular filtration rate (eGFR) < 30 ml/min/1.73 m^2^ and the standard exclusion criteria for CMR (21). CMR was performed on a clinical 1.5T Philips Achieva System (Philips Healthcare, Best, The Netherlands) equipped with a 5-channel cardiac receiver array and a gradient system delivering 80mT/m at 100 mT/m/ms slew rate per physical axis. Eleven patients who underwent AVR were scheduled for a second CMR examination 6 months after surgery. The study protocol was approved by the ethics committee of the canton of Zurich (PB_2016–01944) and conformed to the principles of the Helsinki Declaration. Prior to imaging, written informed consent was obtained from all study subjects. The administration of gadolinium based contrast agents (GBCA) was limited to the patient cohort.

### CMR Data Acquisition

For CMR DTI a diffusion-weighted second-order motion compensated spin-echo sequence was performed with an echo planar imaging readout (22,23) in conjunction with spectral spatial excitation for fat suppression. Basal, midventricular and apical short axis slices were acquired in mid-systole (65% of peak systole) during free breathing with respiratory navigator-based slice tracking (24). Diffusion weighting was encoded along nine and three directions with a b-value of 450 and 100 s/mm^2^, respectively (25).

Additionally, a standard clinical scan protocol was used with balanced steady-state free precession (bSSFP) sequences for the assessment of cardiac volumes and mass. T1 mapping was performed using a Modified Look-Locker Inversion-recovery (MOLLI) sequence with a 5-(3)-3 scheme (26) before, and 20 minutes after a bolus injection of 0.2 mmol/kg GBCA (Gadovist, Bayer (Schweiz) AG, Switzerland). For ECV calculation, the hematocrit was measured in all patients within two hours of CMR. LGE images were acquired with an inversion-recovery sequence approximately ten minutes after the bolus injection. See supplemental Table 1 and supplemental Figure 1 for detailed sequence parameters.

**Figure 1.**
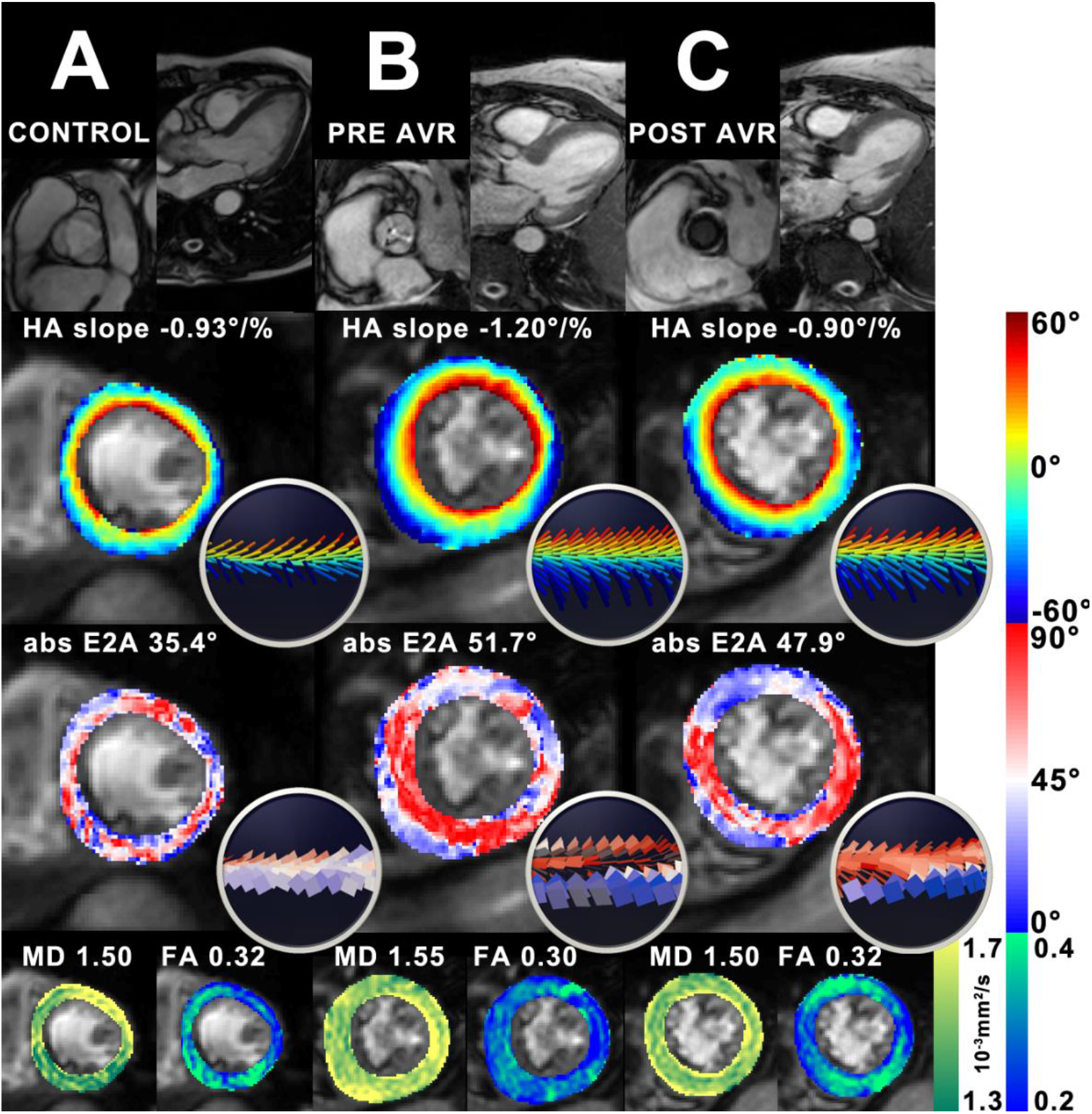
Multiparametric CMR DTI assessment of aortic stenosis and reverse myocardial remodeling after AVR. Panel **A** displays representative DTI maps of a healthy 65-year-old man. In contrast, in panel **B**, an 88-year-old man with severe aortic stenosis (mean gradient 48 mmHg) showed concentric left ventricular hypertrophy with a steeper helix angle (HA) slope and marked elevation of the absolute E2A sheet angle. Mean diffusivity (MD) and fractional anisotropy (FA) were only mildly altered in this case. Panel **C** shows the same patient after transcatheter AVR. HA slope returned to a normal value and there was a 7% reduction in absolute E2A. Similarly, MD and FA showed a trend towards normalisation. The image inlays corresponds to the anterior-septal region.

**Table 1.**
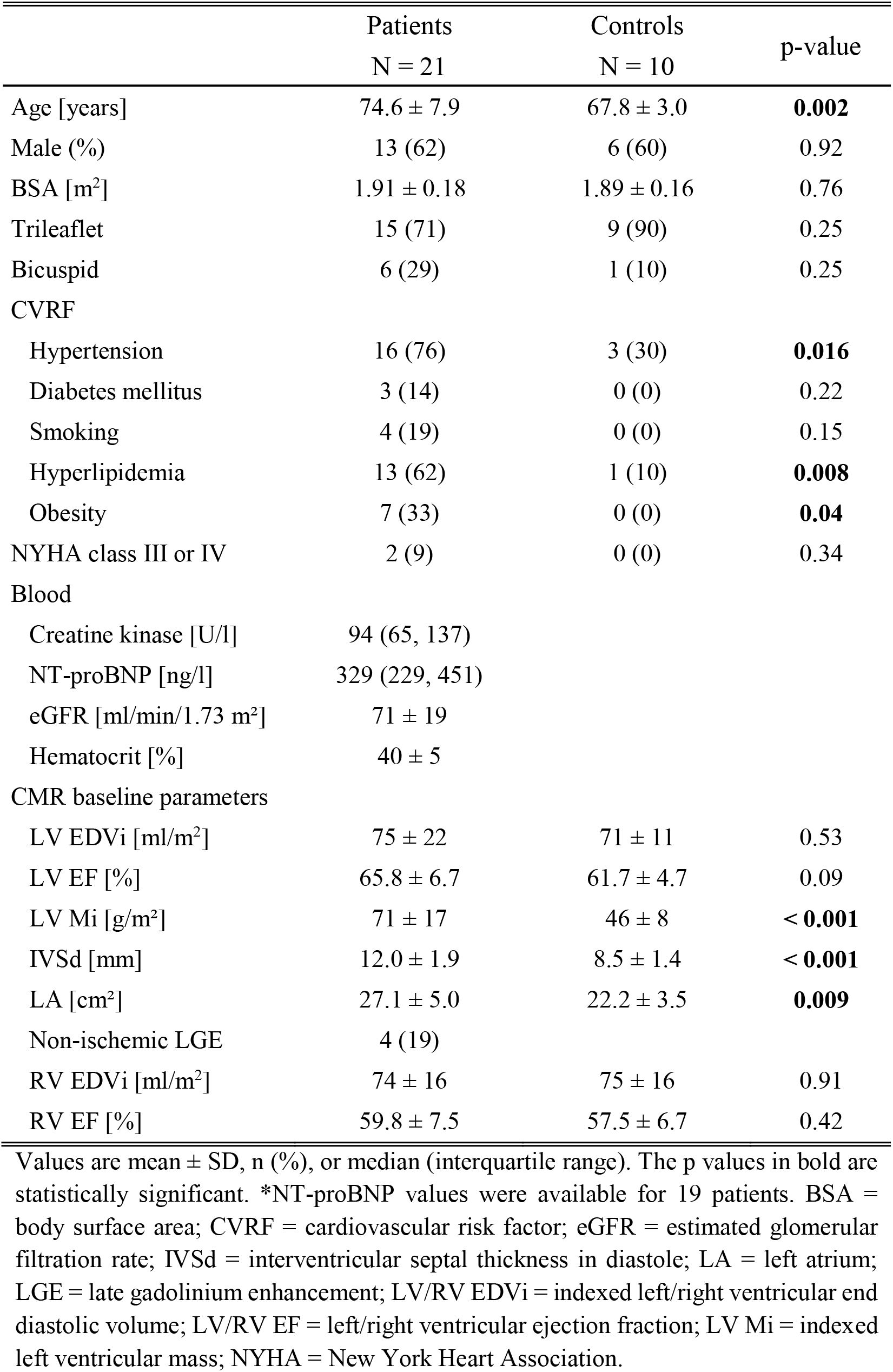
Baseline characteristics of the study cohort

|  | Patients<br>N = 21 | Controls<br>N = 10 | p-value |
| --- | --- | --- | --- |
| Age [years] | 74.6 ± 7.9 | 67.8 ± 3.0 | **0.002** |
| Male (%) | 13 (62) | 6 (60) | 0.92 |
| BSA [m<sup>2</sup>] | 1.91 ± 0.18 | 1.89 ± 0.16 | 0.76 |
| Trileaflet | 15 (71) | 9 (90) | 0.25 |
| Bicuspid | 6 (29) | 1 (10) | 0.25 |
| CVRF |  |  |  |
| Hypertension | 16 (76) | 3 (30) | **0.016** |
| Diabetes mellitus | 3 (14) | 0 (0) | 0.22 |
| Smoking | 4 (19) | 0 (0) | 0.15 |
| Hyperlipidemia | 13 (62) | 1 (10) | **0.008** |
| Obesity | 7 (33) | 0 (0) | **0.04** |
| NYHA class III or IV | 2 (9) | 0 (0) | 0.34 |
| Blood |  |  |  |
| Creatine kinase [U/l] | 94 (65, 137) |  |  |
| NT-proBNP [ng/l] | 329 (229, 451) |  |  |
| eGFR [ml/min/1.73 m<sup>2</sup>] | 71 ± 19 |  |  |
| Hematocrit [%] | 40 ± 5 |  |  |
| CMR baseline parameters |  |  |  |
| LV EDVi [ml/m<sup>2</sup>] | 75 ± 22 | 71 ± 11 | 0.53 |
| LV EF [%] | 65.8 ± 6.7 | 61.7 ± 4.7 | 0.09 |
| LV Mi [g/m<sup>2</sup>] | 71 ± 17 | 46 ± 8 | **< 0.001** |
| IVSd [mm] | 12.0 ± 1.9 | 8.5 ± 1.4 | **< 0.001** |
| LA [cm<sup>2</sup>] | 27.1 ± 5.0 | 22.2 ± 3.5 | **0.009** |
| Non-ischemic LGE | 4 (19) |  |  |
| RV EDVi [ml/m<sup>2</sup>] | 74 ± 16 | 75 ± 16 | 0.91 |
| RV EF [%] | 59.8 ± 7.5 | 57.5 ± 6.7 | 0.42 |

Values are mean ± SD, n (%), or median (interquartile range). The p values in bold are statistically significant. \*NT-proBNP values were available for 19 patients. BSA = body surface area; CVRF = cardiovascular risk factor; eGFR = estimated glomerular filtration rate; IVSd = interventricular septal thickness in diastole; LA = left atrium; LGE = late gadolinium enhancement; LV/RV EDVi = indexed left/right ventricular end diastolic volume; LV/RV EF = left/right ventricular ejection fraction; LV Mi = indexed left ventricular mass; NYHA = New York Heart Association.

### CMR Image Post-Processing and Analysis

CMR image analysis of standard ventricular volumes, mass and function was performed using GTVolume software (GyroTools LLC, Zurich, Switzerland) and manual contouring of the endocardial border at end-diastole and end-systsole and the epicardial border at end-diastole. When appropriate, parameters were indexed to body surface area (BSA) obtained at the day of CMR. LGE was categorized into none, infarct, or non-infarct patterns while patients with signs of ischemic heart disease were excluded from further analysis (n = 1). Patients with only trivial LGE limited to the right ventricular insertion points were considered LGE negative (27).

For the compensation of residual breathing motion, the individual MOLLI and DTI images were registered using a non-rigid groupwise image registration method (28). DTI analysis was performed on mean diffusivity (MD), fractional anisotropy (FA), spherical tensor shape, helix angle (HA) and absolute E2A sheet angle (29,30). The reference longitudinal direction for the calculation of angular diffusion metrics was corrected for the curvature of the LV based on the three DTI slices (31). The transmural HA gradient (HA slope) was determined by the linear regression of the transmural HA distribution, excluding boundary pixels at the endo- and epicardium. The scalar diffusion metrics MD and FA as well as native T1 and ECV, were evaluated in a single septal region of interest on the midventricular short-axis slices for improved reproducibility (32).

### Statistical Analysis

Statistical analyses were conducted using MedCalc software (MedCalc 17.9.7, MedCalc Software Ltd, Ostend, Belgium). Continuous variables are expressed as mean±SD or as median and interquartile range (IQR) for not normally distributed data. Categorical variables are expressed as counts and percent. Normal distribution of data was confirmed using the Shapiro-Wilk test. Differences between AS patients and controls were assessed with the unpaired Student t test, Mann-Whitney-U test, or χ^2^ test as appropriate. Changes in parameters after AVR were assessed using the paired Student t test or Wilcoxon test. To determine statistical associations between parameters, Pearson correlation analyses were performed. A p-value smaller than 0.05 was considered statistically significant.

## Results

### Patient Characteristics

Data from one of 22 AS patients was excluded due to co-existing ischemic heart disease. Hence, a total of 21 AS patients (74.6±7.9 years, 62% male) were included in the final analysis. The baseline characteristics of AS patients and control subjects are summarized in Table 1. The sex-matched senior control subjects were on average 6.8 years younger than the AS patients but had similar BSA characteristics. As expected, cardiovascular risk factors were more common in the AS cohort, in particular hypertension (76%), hyperlipidaemia (62%) and obesity (33%). Bicuspid aortic valves were present in six AS patients and one control subject (p = 0.25). The most common symptom in the AS group was mild dyspnea (NYHA II, 62%), 33% had chest pain, 14% vertigo or syncope and 10% had severe dyspnea (NYHA III). Median NT-proBNP in the AS group was 329 ng/l while 5 patients (24%) had NT-proBNP values above their sex-and age-specific reference value. AS patients exhibited significantly elevated LV mass (p< 0.001), thickness of the interventricular septum (p< 0.001) and left atrial size (p = 0.009). Four AS patients had non-infarct patters of LGE. LV and RV volumes and ejection fraction were not significantly different between the groups.

### DTI Parameters in Aortic Stenosis vs Controls

Representative diffusion and structural parameter maps of an AS patient and a control subject are shown in Figure 1A & 1B. In AS patients, the DTI diffusion parameters showed significantly elevated MD (AS 1.56±0.07 vs controls 1.47±0.06 10^−3^ mm^2^/s, p = 0.002) and reduced FA (AS 0.29±0.03 vs controls 0.35±0.03, p< 0.001). In accordance with reduced FA, there was increased tensor sphericity in the AS cohort (AS 0.58±0.03 vs controls 0.53±0.02, p< 0.001) indicating more diffusion orthogonal to the predominant direction. Regarding the DTI orientation parameters, AS patients exhibited a marked elevation of HA slope (AS –1.13±0.11 vs controls –0.95±0.12°/%depth, p< 0.001). Moreover, there was a significantly increased absolute E2A sheet angle (AS 45.7±6.1 vs 35.0±7.9°, p< 0.001) in the AS group. Absolute E2A sheet angle showed a highly significant correlation with the interventricular septum thickness in diastole (IVSd) (r = 0.69; p< 0.0001) over the entire study population. Table 2 contains the comparison of all parameters between AS patients and controls.

**Table 2.**
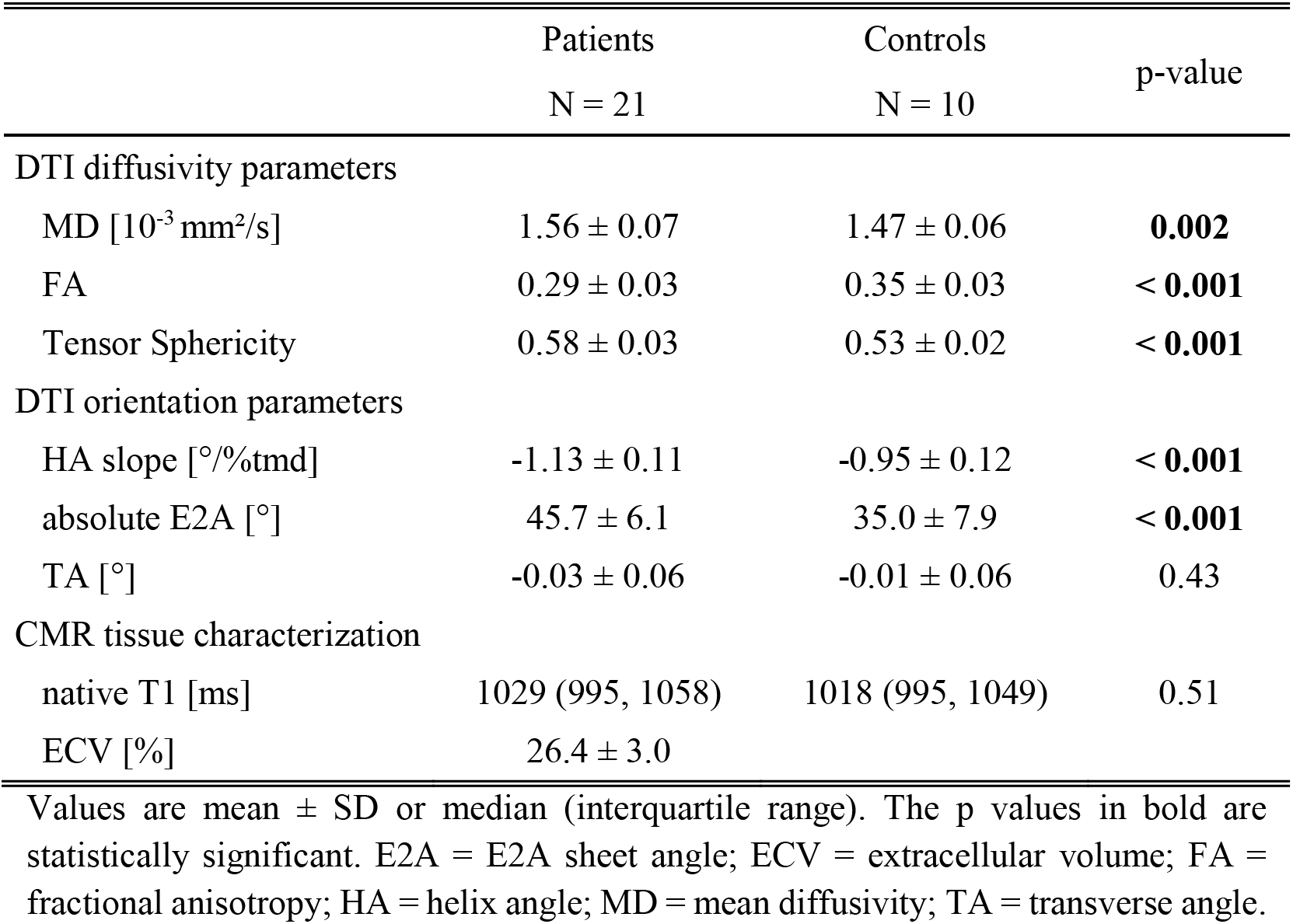
Comparison of CMR tissue characterization, diffusion and structural parameters between aortic stenosis and healthy controls

|  | Patients<br>N = 21 | Controls<br>N = 10 | p-value |
| --- | --- | --- | --- |
| DTI diffusivity parameters |  |  |  |
| MD [$10^{-3}$ mm<sup>2</sup>/s] | 1.56 ± 0.07 | 1.47 ± 0.06 | **0.002** |
| FA | 0.29 ± 0.03 | 0.35 ± 0.03 | **< 0.001** |
| Tensor Sphericity | 0.58 ± 0.03 | 0.53 ± 0.02 | **< 0.001** |
| DTI orientation parameters |  |  |  |
| HA slope [°/%tmd] | -1.13 ± 0.11 | -0.95 ± 0.12 | **< 0.001** |
| absolute E2A [°] | 45.7 ± 6.1 | 35.0 ± 7.9 | **< 0.001** |
| TA [°] | -0.03 ± 0.06 | -0.01 ± 0.06 | 0.43 |
| CMR tissue characterization |  |  |  |
| native T1 [ms] | 1029 (995, 1058) | 1018 (995, 1049) | 0.51 |
| ECV [%] | 26.4 ± 3.0 |  |  |

Values are mean ± SD or median (interquartile range). The p values in bold are statistically significant. E2A = E2A sheet angle; ECV = extracellular volume; FA = fractional anisotropy; HA = helix angle; MD = mean diffusivity; TA = transverse angle.

### Reversibility of Structural and Diffusion Alterations Following AVR

After a median follow-up of 6 months (IQR: 5.0 to 6.9 months) post AVR, 11 patients underwent a second CMR (4 surgical AVR, 7 transcatheter AVR). Figure 1B & 1C compare DTI parameter maps of a patient before and after transcatheter AVR. There was a significant reduction of LV Mi (p = 0.007) and IVSd (p = 0.001) after AVR, while LV EDVi and EF remained unchanged. CMR DTI diffusivity analysis showed a significant regression towards normal values for MD (p = 0.014), FA(p = 0.011) and tensor sphericity (p = 0.011) (Figure 2A & 2B). Similarly, the DTI orientation parameters experienced a significant normalization after AVR, in particular the HA slope (p< 0.001) reached the level of the healthy control group (Figure 2C). Also absolute E2A sheet angle decreased significantly after AVR (p = 0.003) but remained on average 7° above the level of healthy controls (Figure 2D). Table 3 summarises the findings in AS patients pre and post AVR.

**Figure 2.**
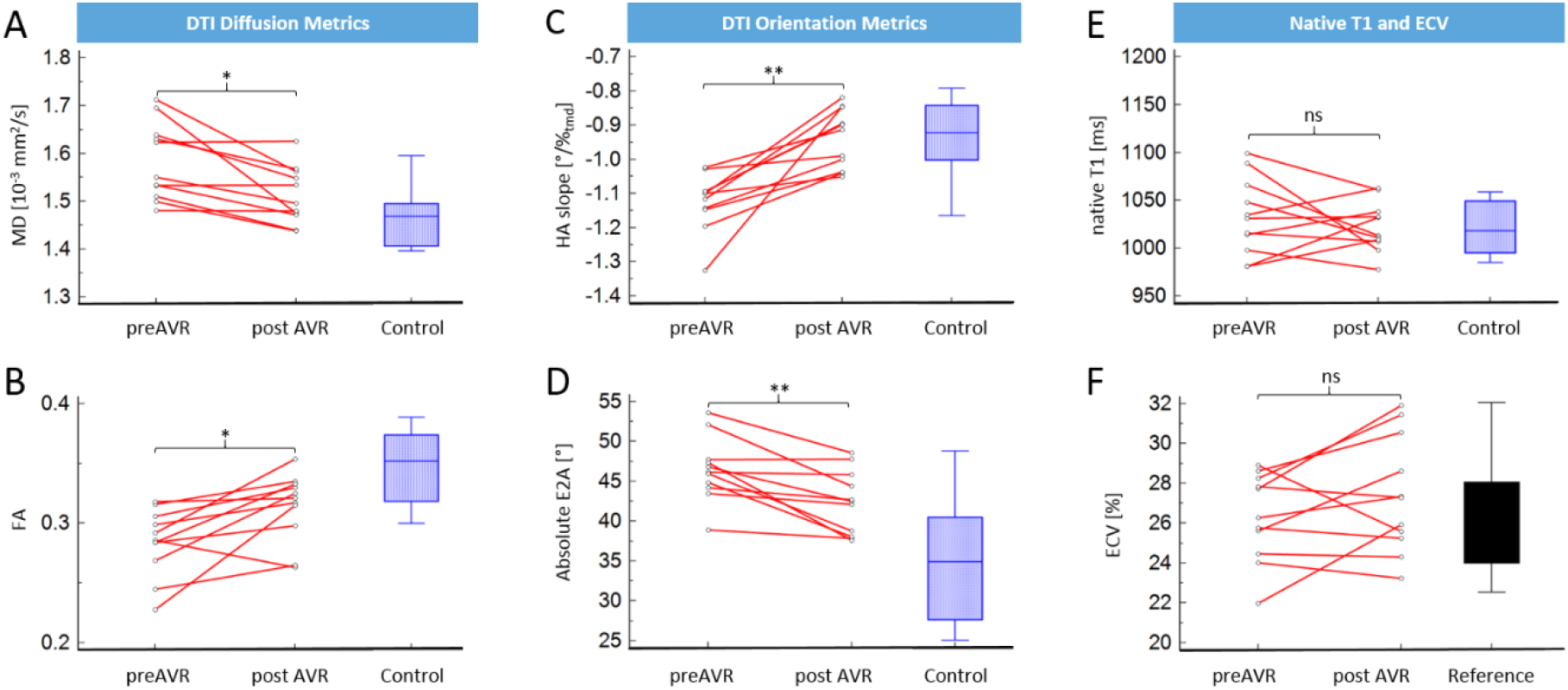
Dynamic of DTI diffusion metrics and DTI orientation metrics compared to native T1 and ECV 6 months after AVR. At 6 months post AVR, there was a 4% reduction in mean diffusivity (**A**) and a 9% increase in fractional anisotropy (**B**). The helix angle slope returned to normal level in the majority of cases with a relative increase of 16% (**C**). The absolute E2A sheet angle reduced by 9% post AVR but was on average still 7° higher than healthy controls (**D**). Native T1 showed a non-significant trend to lower values after AVR (**E**) and the extracellular volume fraction increased non-significantly (**F**).

**Table 3.** Changes after aortic valve replacement (N = 11)

|  | pre AVR | post AVR | p-value |
| --- | --- | --- | --- |
| LV EDVi [ml/m<sup>2</sup>] | 71 ± 13 | 77 ± 20 | 0.27 |
| LV EF [%] | 66.1 ± 5.2 | 66.9 ± 4.5 | 0.69 |
| LV Mi [g/m<sup>2</sup>] | 69 ± 14 | 59 ± 8 | **0.007** |
| IVSd [mm] | 12.0 ± 2.0 | 10.2 ± 1.4 | **0.001** |
| LA [cm<sup>2</sup>] | 26.7 ± 5.8 | 26.9 ± 6.3 | 0.85 |
| Blood |  |  |  |
| Hct [%] | 38.8 ± 6.4 | 38.9 ± 6.4 | 0.90 |
| CMR tissue characterization |  |  |  |
| native T1 [ms] | 1033 ± 40 | 1022 ± 26 | 0.43 |
| ECV [%] | 26.3 ± 2.2 | 27.4 ± 2.9 | 0.16 |
| DTI diffusivity parameters |  |  |  |
| MD [10<sup>-3</sup> mm<sup>2</sup>/s] | 1.58 ± 0.08 | 1.52 ± 0.06 | **0.014** |
| FA | 0.28 ± 0.03 | 0.31 ± 0.03 | **0.011** |
| Sphericity | 0.58 ± 0.03 | 0.56 ± 0.03 | **0.011** |
| DTI orientation parameters |  |  |  |
| HA slope [°/%tmd] | -1.12 ± 0.08 | -0.94 ± 0.08 | **< 0.001** |
| absolute E2A [°] | 46.5 ± 4.0 | 42.4 ± 4.0 | **0.003** |
| TA [°] | -0.02 ± 0.05 | -0.01 ± 0.07 | 0.45 |

Values are mean ± SD. The p values in bold are statistically significant. E2A = E2A sheet angle; ECV = extracellular volume; FA = fractional anisotropy; HA = helix angle; Hct = haematocrit; IVSd = interventricular septal thickness in diastole; LA = left atrium; LV EDVi = indexed left ventricular end diastolic volume; LV EF = left ventricular ejection fraction; LV Mi = indexed left ventricular mass; MD = mean diffusivity; TA = transverse angle; TA SD = standard deviation of TA.

### Comparison of Native T1 and ECV with DTI Parameters in AS

Native T1 mapping was not significantly different between AS patients and healthy controls (AS 1029 ms [IQR: 995, 1058] vs controls 1018 ms [IQR: 995, 1049], p = 0.51). Also, there was no significant difference between pre and post AVR native T1 (pre 1033±40 vs post 1022±26 ms, p = 0.43, Figure 2E) and between pre and post AVR ECV (pre 26.3±2.2 vs post 27.4±2.9 %, p = 0.16 Figure 2F). Pre and post ECV stayed within the reference range of healthy controls according to (33). In the AS patient group, none of the fibrosis marker i.e. native T1 and ECV was correlated with the DTI diffusion parameters MD or FA. After AVR, however, native T1 was significantly correlated with both MD (r = 0.71, p = 0.014) and FA (r = –0.60, p = 0.050). ECV and MD show a non-significant trend towards a positive correlation after AVR (r = 0.44, p = 0.17) while there was a non-significant negative correlation before AVR (r = –0.20, p = 0.40).

## Discussion

In patients with severe aortic stenosis, CMR DTI was found to provide a set of parameters to characterize myocardial response to pressure overload and to monitor reverse remodeling after AVR (Central Illustration).

We show that AS is accompanied by an increase in free myocardial diffusion which can be quantified by significantly increased MD and reduced FA. Probably due to an early referral practice, the AS cohort in the present study did not show extensive myocardial fibrosis with a low number of LGE positive cases, no significant elevation of native T1 and a relatively normal ECV compared with previous studies in AS patients (9,14). Therefore, the data suggests that the MD and FA alterations in this study represent predominantly increased intracellular diffusion caused by cellular hypertrophy(34). This concept is supported by increased tensor sphericity of diffusion, which indicates that the additional diffusion occurs predominantly perpendicular to the myocyte orientation (primary diffusion eigenvector) as expected in cellular hypertrophy. In cardiac diseases with primarily extracellular pathology, such as cardiac amyloidosis, where extracellular accumulation of misfolded proteins is a key pathogenic factor, MD and FA exhibit good correlation with ECV (18). The lack of correlation between MD or FA and measures of myocardial fibrosis in the AS cohort indicates that the DTI diffusion metrics provide complementary information in particular regarding the cellular myocardial response to pressure overload that is not captured by ECV or native T1 alone. Combined with previous data showing deranged MD and FA values in various cardiomyopathies, including HCM, DCM or myocardial infarction this study suggests that DTI diffusion measures are capable of identifying pathological states of the extracellular matrix as well as of the myocytes. Potential prognostic value of FA has recently been shown in HCM patients where low FA was associated with ventricular arrhythmias even after accounting for LGE and ECV (35). This finding underlines the independent contribution of cellular disarray and hypertrophy to adverse outcome. Since cellular hypertrophy is the primary early myocardial response not only in AS but also in hypertension, in-vivo characterization of cardiomyocyte abnormalities due to pressure overload may provide novel insights into the pathophysiology of the most common cardiovascular risk factor. From a risk stratification perspective, it would be of particular interest if DTI measures are able to differentiate physiological hypertrophy as found in athlete hearts from early adverse remodeling caused by diseases with increased LV afterload.

Besides changes in myocardial diffusion, our investigation also revealed marked microstructural alterations in AS patients. Similar to previous reports in HCM and amyloidosis (18,19), the E2A sheet angle was elevated in AS patients. The significant correlation between the thickness of the septal wall and E2A substantiates the hypothesis that not only cellular hypertrophy or ECV expansion, but also exaggerated tilting of the laminar sheetlet orientation contributes to myocardial thickening. This mechanism appears to be a common final path of multiple cardiomyopathies with hypertrophic phenotype independent of the initial pathologic stimulus.

In contrast, the HA slope appears to vary significantly with different diseases of hypertrophic phenotype. As shown in this study, the HA slope is significantly steeper in AS patients than in controls. This local adaption of myocyte orientation may allow for the increased torsion and twist seen in patients with pressure overload hypertrophy due to aortic stenosis (36). Previous computational models of myocardial structure predicted adaption of the helix angle distribution to minimizie myocardial fiber strain (37,38). While elevated in AS, the HA slope was found to be significantly reduced in cardiac amyloidosis (18). The apparent variation of the HA slope depending on the pathologic stimulus that lead to hypertrophy makes this parameter particularly interesting for phenotyping diseased myocardium.

After AVR, the myocardial diffusion parameters MD and FA experienced a clear trend towards normalization. This is of particular interest, since native T1 did not show a significant alteration due to AVR and ECV exhibited even a trend to higher values. Those results for ECV and native T1 resemble the findings of a previous study in 116 patients, where the ECV expansion after AVR has been attributed to a different timelines for the regression of the cellular and the extracellular compartment with normalization of the extracellular matrix being slower (12). While cellular regression can be expected approx. 6 months after reduction of afterload, elevated ECV persists for at least 12 months (12,39). In good agreement with that observation, MD and FA in our study did not reach the level of healthy controls, indicating that the observed normalization can be attributed to the regression of cellular hypertrophy and disarray while the remaining alteration compared to controls reflects the increased diffusion in the extracellular compartment.

Similarly, the DTI orientation parameters showed significant regression after AVR. HA slope reached the level of healthy controls at 6 months which indicates that the adaption of myocyte orientation to reduced afterload is a dynamic process. Also, E2A sheet angle decreased by 9% but remained elevated compared to controls, which is in line with residual global LV hypertrophy found 6 months after AVR in this and previous studies (40).

The DTI parameter exhibited larger relative changes in response to AS and AVR when compared to the conventional tissue characterization measures native T1 and ECV (Figure 4). Even when significant changes in native T1 between healthy volunteers and AS subjects were observed in larger studies (10), the relative change was still < 5%. Similarly, ECV exhibited only a small increase in AS and showed further expansion after AVR. In contrast, all DTI parameters showed changes > 5% in response to pressure overload and regression after AVR. Therefore, DTI may open a new window to phenotype myocardial adaption in AS beyond fibrosis by integrating alterations of the extracellular and cellular myocardial compartments. Future research is warranted to investigate the potential of CMR DTI as imaging biomarker for further optimization of treatment planning.

**Figure 3.**
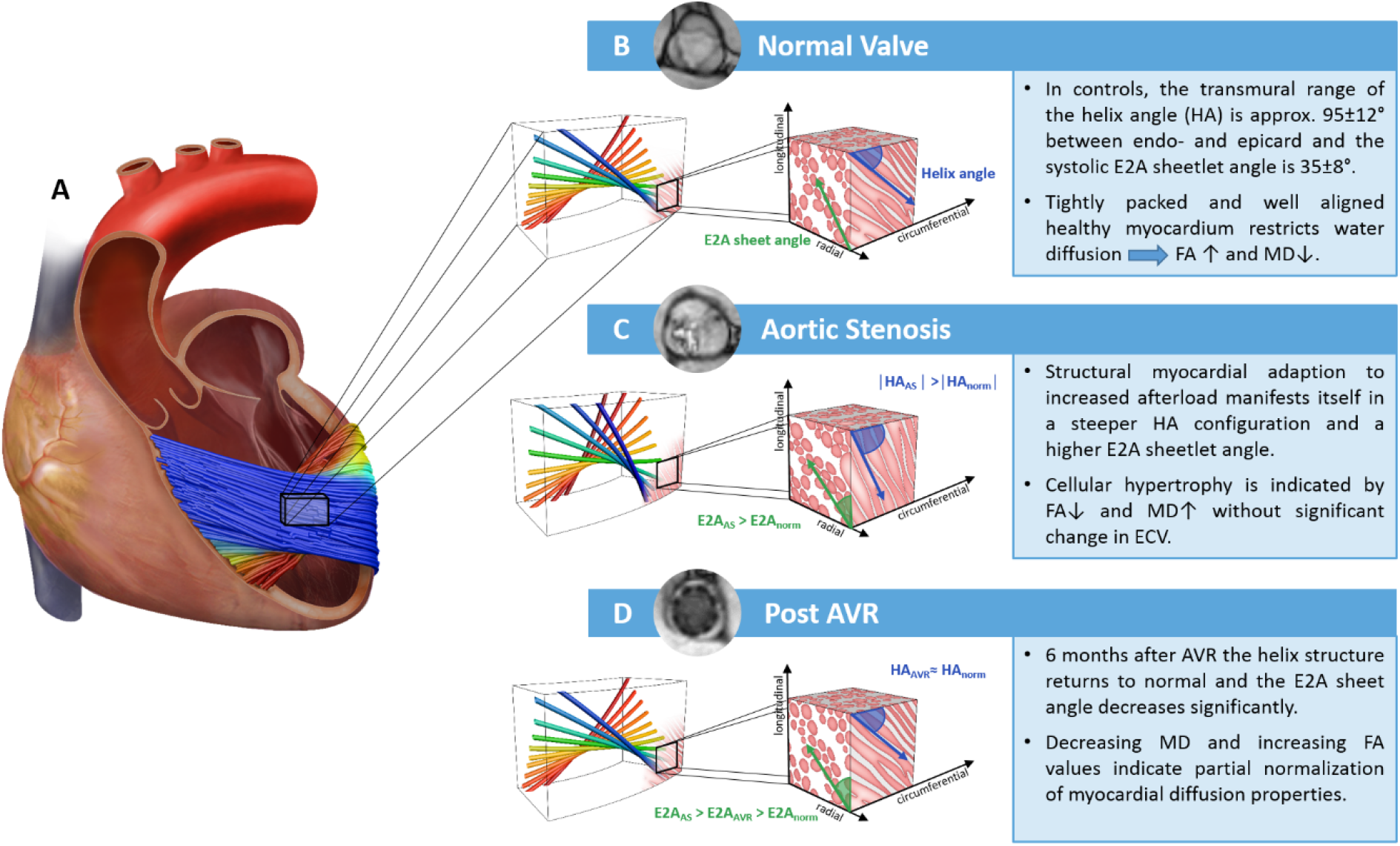
Central Illustration. (**A**) 3-dimensional tractogram obtained from in-vivo CMR diffusion tensor imaging overlaid on a symbolic human heart shows the helical structure of the myocardium. (**B**) Zoom on a schematic diagram of the helical structure in a normal heart on the left. A symbolic myocardial block on the right shows the helix angle (HA) on an epicardial plane and the sheetlets, which are closely packed cardiomyocytes, in an orthogonal cutting plane. (**C**) Aortic stenosis leads to a steeper distribution of the helix angle and a higher sheetlet angle (E2A). (**D**) After aortic valve replacement, the E2A regresses significantly and the helix angle returns to normal.

**Figure 4.**
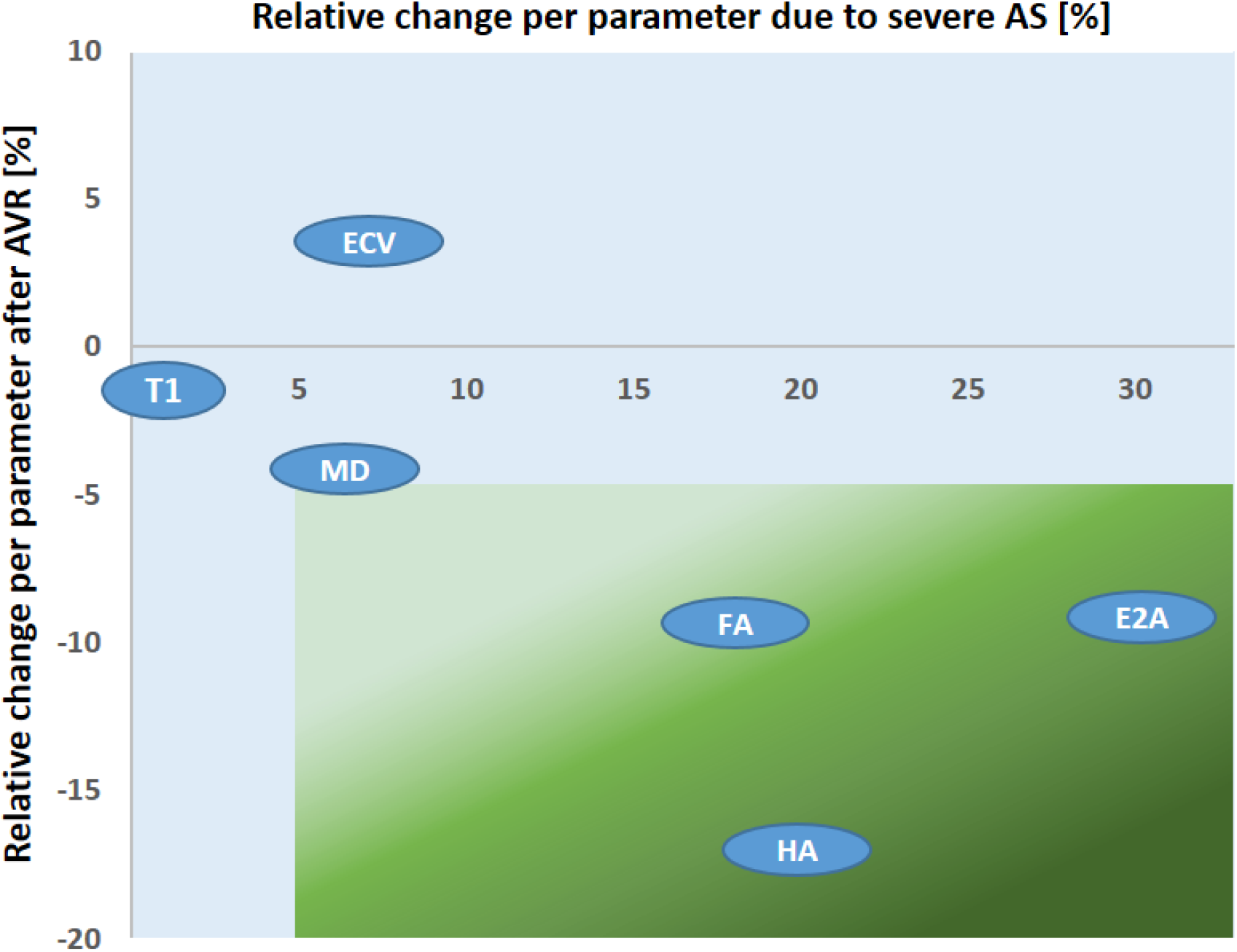
Relative change of the tissue characterisation parameters due to AS and after AVR. This plot shows parameters with at least 5% relative change due to AS and after AVR in the right lower area (green). Fractional anisotropy (FA), absolute E2A sheet angle (E2A) and the helix angle (HA) slope exhibit marked changes due to pressure overload, that are largely reversible after AVR. The mean diffusivity (MD) shows borderline changes of approx. 5% in both directions. Native T1, in contrast, displays only minimal relative changes and ECV is the only parameter that shows no reversibility, but increases after AVR.

### Limitations

Besides fibrosis and cellular hypertrophy, the pathologic response to AS may also include myocardial edema or vasodilation which were not accounted for in this study. However, those effects are believed to be limited since no patient exhibited myocardial edema in T2-weighted sequences and capillary density is reduced in pressure overload hypertrophy (41). Also, reverse remodeling in AS is not finished after 6 months and additional changes of DTI parameters may occur at a later stage. However, the timeframe for post-interventional imaging was chosen to reflect the end of cellular reverse remodeling (39) while extracellular regression is known to continue for years. Other limitations of the present study include the investigation of only the systolic configuration and small sample size in this single center approach.

### Conclusion

CMR DTI has identified novel characteristic alterations of myocardial diffusion properties and of cardiomyocyte orientation caused by aortic stenosis. Following AVR, both diffusion abnormalities and structural changes regress and show higher plasticity when compared to native T1 or ECV. Therefore, CMR DTI provides a new set of parameters that may help the understanding of the reversibility of AS-induced myocardial damage and to add information for optimizing AVR timing before irreversible adverse remodeling occurres.

## Data Availability

Data availability is subject to reasonable request and ethical guidelines

## Abbreviations List

AS: aortic stenosis
AVR: aortic valve replacement
BSA: body surface area
bSSFP: balanced steady-state free precession
CMR: cardiovascular magnetic resonance
DCM: dilated cardiomyopathy
DTI: diffusion tensor imaging
E2A: absolute E2A sheet angle
ECV: extracellular volume
FA: fractional anisotropy
GBCA: gadolinium based contrast agent
HA: helix angle
HCM: hypertrophic cardiomyopathy
IVSd: interventricular septum thickness in diastole
LGE: late gadolinium enhancement
LV EDV: left ventricular end-diastolic volume
LV EF: left ventricular ejection fraction
LV Mi: left ventricular indexed mass
MD: mean diffusivity
MOLLI: modified look-locker inversion recovery
RV EDV: right ventricular end-diastolic volume
RV EF: right-ventricular ejection fraction

## Acknowledgments

This work is supported by the Swiss National Science Foundation: CR23I3_166485 and PZ00P2_174144.

## Supporting Information

**Supp Table 1.** imaging parameters.

|  | Cine bSSFP | LGE | T1 | DTI |
| --- | --- | --- | --- | --- |
| spatial resolution [mm<sup>2</sup>] | 1.8×1.8 | 1.6×1.6 | 2.4×2.4 | 2.6×2.6 |
| slice thickness | 8 | 10 | 10 | 8 |
| encoding | 2D | 3D | 2D | 2D slice cycling |
| no heart phases | 24 | 1 | 1 | 1 |
| No slices | 12-14 | 10 | 3 | 3 |
| TR/TE [ms] | 3.4/1.7 | 3.6/1.7 | 2.3/1.0 | 3000/76 |
| undersampling factor | 2 | 2 | 2 | 1 |

**Supp Figure 1.**
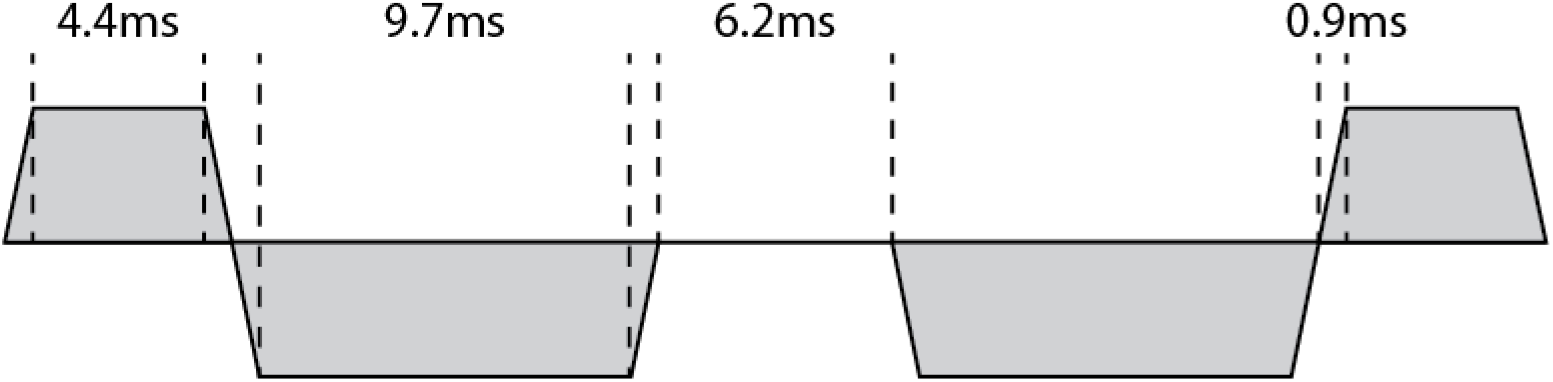
timing of second order motion compensated diffusion encoding gradient waveform.

